# Machine Learning Models Predict the Emergence of Depression in Argentinean College Students during Periods of COVID-19 Quarantine

**DOI:** 10.1101/2024.01.25.24301772

**Authors:** Lorena Cecilia López Steinmetz, Margarita Sison, Rustam Zhumagambetov, Juan Carlos Godoy, Stefan Haufe

## Abstract

**INTRODUCTION:** The COVID-19 pandemic has exacerbated mental health challenges, particularly depression among college students. Detecting at-risk students early is crucial but remains challenging, particularly in developing countries. Utilizing data-driven predictive models presents a viable solution to address this pressing need. **AIMS: 1)** To develop and compare machine learning (ML) models for predicting depression in Argentinean students during the pandemic. **2)** To assess the performance of classification and regression models using appropriate metrics. **3)** To identify key features driving depression prediction.

**METHODS:** A longitudinal dataset (N = 1492 college students) captured T1 and T2 measurements during the Argentinean COVID-19 quarantine. ML models, including linear logistic regression classifiers/ridge regression (LogReg/RR), random forest classifiers/regressors, and support vector machines/regressors (SVM/SVR), are employed. Assessed features encompass depression and anxiety scores (at T1), mental disorder/suicidal behavior history, quarantine sub-period information, sex, and age. For classification, models’ performance on test data is evaluated using Area Under the Precision-Recall Curve (AUPRC), Area Under the Receiver Operating Characteristic curve, Balanced Accuracy, F1 score, and Brier loss. For regression, R-squared (R2), Mean Absolute Error, and Mean Squared Error are assessed. Univariate analyses are conducted to assess the predictive strength of each individual feature with respect to the target variable. The performance of multi- vs univariate models is compared using the mean AUPRC score for classifiers and R2 score for regressors.

**RESULTS:** The highest performance is achieved by SVM and LogReg (e.g., AUPRC: 0.76, 95% CI: 0.69, 0.81) and SVR and RR models (e.g., R2 for SVR and RR: 0.56, 95% CI: 0.45, 0.64 and 0.45, 0.63, respectively). Univariate models, particularly LogReg and SVM using depression (AUPRC: 0.72, 95% CI: 0.64, 0.79) or anxiety scores (AUPRC: 0.71, 95% CI: 0.64, 0.78) and RR using depression scores (R2: 0.48, 95% CI: 0.39, 0.57) exhibit performance levels close to those of the multivariate models, which include all features.

**DISCUSSION:** These findings highlight the relevance of pre-existing depression and anxiety conditions in predicting depression during quarantine, underscoring their comorbidity. ML models, particularly SVM/SVR and LogReg/RR, demonstrate potential in timely detection of at-risk students, enabling preventive measures for improved mental health outcomes.

## INTRODUCTION

The COVID-19 pandemic has presented significant global challenges to mental health, particularly among college students. The sudden transition to remote learning, social isolation, and economic instability has exacerbated mental health issues, with depression becoming a prevalent concern for this population (Li et al., 2021; Wang et al., 2021). Depression can negatively impact academic performance, social relationships, and overall quality of life. Therefore, early identification of depression and its risk factors is crucial for timely interventions and prevention of further negative outcomes. However, identifying individuals who may be at risk of developing depression remains a significant challenge in this field.

The literature consistently identifies key psychological and demographic variables linked to or predicting depression. Anxiety often emerges as a predictor for depression (American Psychiatric Association, 2000; Horn & Wuyek, 2010), notably among college students (Cassady et al., 2019). Moreover, histories of diagnosed mental disorders (Sheldon et al., 2021) and suicidal behavior (Kisch et al., 2005) closely correlate with college student depression. While sex (Piccinelli & Wilkinson, 2000) and age (Nolen-Hoeksema & Aldao, 2011) differences in depression are well-documented, sex-based variations in depression may display age-specific trends (Jorm, 1987). Amid COVID-19 lockdowns, mental health, particularly depression, was anticipated to deteriorate with extended measures, impacting well-being during and after implementation (Brooks et al., 2020). Yet, achieving a comprehensive integration of these factors into accurate predictive models for depression remains incomplete and challenging.

Existing literature on depression has extensively analyzed variability between groups and within individuals (e.g., Jacobson et al., 2017). Mixed-effects modeling (MEM), or hierarchical linear modeling, is a widely used statistical approach that allows for estimation of both fixed and random effects, accounting for variability between groups and within individuals. MEM has a major strength in its ability to identify individual and group-level effects, as well as interactions between them, which is particularly relevant in fields like psychology where individual differences are of interest.

However, MEM is primarily designed for inference rather than prediction, and offers limited flexibility for the handling of high-dimensional data, compromising their predictive accuracy compared to machine learning (ML) models. ML models are well-suited for prediction tasks, handling complex relationships between variables and relying less on strong data assumptions (Hastie et al., 2009; James et al., 2017; Kuhn et al., 2013; Murphy, 2012). MEM, in particular, relies on assumptions about predictor-response relationships (i.e., linearity), normality, and independence of residuals, which can impact their predictive accuracy for new data. Conversely, ML is a data-driven approach that uses algorithms to identify patterns in the data for prediction. However, ML models require extensive data for training and can be prone to overfitting with too complex models or small datasets. Therefore, while ML has emerged as an alternative or complementary approach to traditional statistics in mental health research, it is an open challenge to leverage ML for predicting individuals at risk for depression.

In the field of mental health assessment and ML applications, the challenge of diagnosis prediction can be tackled using a dual methodology, encompassing both classification and regression approaches. In the context of binary classification, the goal is to categorize individuals into two groups: those with depression and those without. This approach yields a straightforward determination of depression presence, aiding in identifying individuals requiring additional evaluation. Conversely, in the context of regression, the aim is to estimate or forecast numerical depression scores. This approach offers a continuous prediction of depression severity, affording a deeper insight into the condition and potentially enabling personalized treatment strategies.

In a prior study, we utilized MEM to investigate the aforementioned key psychological and demographic factors linked to or predictive of depression in college students during Argentina’s COVID-19 pandemic quarantine, which was one of the most rigorous and extended lockdowns globally (López Steinmetz et al., 2023). In the current study, we seek to reassess the same dataset using ML algorithms to evaluate their potential as an alternative or complementary technique to MEM for predicting depression diagnosis. Through this, we hope to enhance predictive precision and effectively identify individuals with depression, while gaining deeper insights into the intricate interplay of factors contributing to depression during pandemics. Furthermore, we expect to determine whether the key factors commonly acknowledged as depression predictors remain as significant input features when utilizing ML algorithms for data analysis, in contrast to other widely used statistical methodologies in psychology. The objectives of this study are: **1)** To develop and compare various ML models, including linear logistic regression classifiers/ridge regression, random forest classifiers/regressors, and support vector machines/support vector regression models, to predict depression in college students during the pandemic based on psychological inventory scores, basic clinical information, quarantine sub-period information, and demographics as features. **2)** To assess the performance of classification and regression models using appropriate metrics. **3)** To identify the key features that drive the prediction of depression by applying univariate methods.

A recent review following the PRISMA guidelines identified only 33 peer-reviewed studies in the domain of utilizing ML algorithms for predicting mental health diagnoses, with only 31% specifically focusing on depression/anxiety disorders. Additionally, most studies have been conducted on clinical samples, primarily consisting of adults or older patients (Iyortsuun et al., 2023). It is worth noting that many of these studies rely on unavailable facilities and resources for diagnosis, particularly in developing countries, such as MRI or blood samples. Therefore, one novel aspect of this study is its focus on exploring the potential of ML algorithms to identify risk factors for depression, during a pandemic, in a large longitudinal sample of quarantined and apparently healthy college students from a developing country. By doing so, it aims to provide further insights into this important area of research by enhancing the understanding of key determinants in depression detection and may have significant implications for the development of more effective screening tools and interventions for depression in college students during pandemics.

## MATERIALS AND METHODS

### Research Design and Dataset

This study employs a longitudinal dataset featuring two-repeated measurements in college students during the Argentinean COVID-19 quarantine period. The first measurement (T1) was taken across various quarantine sub-periods, each characterized by varying levels of restrictions, and spanning up to 106 days. The follow-up measurement (T2) occurred one month later (as depicted in **Figure S1**).

The choice of this longitudinal research design arises from its inherent capability to capture dynamic changes over time, a crucial aspect when investigating the effects of a rapidly evolving event like the COVID-19 pandemic. This approach transcends mere cross-sectional snapshots, providing a more thorough understanding of the intricate interplay between psychological states and external factors amid a quarantine. By incorporating two measurements at distinct time points, it becomes possible to discern short-term fluctuations and potential enduring effects. The T1 measurement establishes a baseline and assesses the immediate effects of quarantine sub-periods lasting up to 106 days. The T2 measurement, conducted a month later, offers insights into the persistence or evolution of mental health patterns, shedding light on potential longer-term consequences. This temporal depth captures nuances that a single-time assessment might overlook, providing a more nuanced portrayal of the challenges college students faced during this unprecedented period.

The dataset comprises responses from 1492 college students who completed an online survey during the mandatory restrictive quarantine. It encompasses assessments of depression, anxiety-trait, basic clinical information, demographics, and quarantine sub-periods. This dataset corresponds to the one utilized in our previous study (López Steinmetz et al., 2023). Further information on the research design and data collection can be found there and in López Steinmetz (2023).

### Input Features

The prediction models were developed using the following input features: age, sex (female, male), history of diagnosed mental disorder (absent, present), history of suicidal attempt and/or ideation (absent, present), depression scores from T1 using the Argentinean validation (Brenlla & Rodríguez, 2006) of the Beck Depression Inventory (BDI) (Beck et al., 1996), anxiety-trait scores from T1 using the Spanish version of the State-Trait Anxiety Inventory (Spielberger et al., 1983), and three quarantine sub-periods to which participants’ responses were chronologically assigned. These sub-periods were categorized according to decreasing levels of COVID-related restrictions over the 106-day duration, and participants were classified into one of three quarantine sub-periods based on the date of their response for measurement T1 (**Figure S1**).

### Target Variable

The target variable, depression, was assessed using the Argentinean validation (Brenlla & Rodríguez, 2006) of the BDI (Beck et al., 1996). For the classification task, depression scores from T2 were labeled as either according to the absence or presence of clinically relevant levels of depression, respectively. The standardized cut-off score of >20, indicative of depression presence in non-clinical populations (Kendall et al., 1987), was employed. For the regression task, raw depression scores were used as a continuous variable.

### Software

The analysis was performed using Python software (version 3.11.1) and relevant libraries, including scikit-learn (Pedregosa et al., 2011), pandas (McKinney, 2010), and numpy (Walt et al., 2011), among others.

### Data analysis

#### - Preprocessing

The dataset does not contain missing data. Before performing classification and regression tasks, the data were randomly divided into a training (75%) and a test (25%) set. In the classification task, the proportion of negative to positive labels was maintained in both sets. Continuous features such as depression and anxiety scores from T1 and age were scaled using a quantile transformation, specifically, we used the QuantileTransformer method (Pedregosa et al., 2011) setting the output distribution as normal, to reduce skewness and enhance model performance. Principal component analysis (PCA) was applied for dimensionality reduction. PCA was set to retain the number of components essential to account for 95% of the variance present in the original data. Consequently, a total of 3 components were retained. Both feature scaling and dimensionality reduction were fitted on the training set and subsequently applied to both sets. The target variable in the regression task was scaled using the same quantile transform function.

#### - Hyperparameter tuning in the classification and regression tasks

Both in the classification task and in the regression task hyperparameter tuning was conducted using the GridSearchCV function of the scikit-learn library (Pedregosa et al., 2011) on the training set for each model. Stratified 10-fold cross-validation, which is typically used when dealing with imbalanced datasets, was applied for robust evaluation during hyperparameter tuning, that is, the training data were split into ten stratified subsets, where in each fold one of the subsets served as an inner validation set and nine as an inner training set. Three ML models for the classification task and three ML models for the regression task with different hyperparameter choices were optimized on each inner training set and tested on each inner validation set. In the classification task, the Average Precision (AP) score was used as the evaluation metric to select model hyperparameters within the inner cross-validation. For the regression task, the R-squared (R2) score was employed as the evaluation metric for hyperparameter selection within the inner cross-validation process. The state of the random number generator was set to 0 for reproducibility in all cases. The final model for each ML model was then trained on the entire training set using the selected hyperparameters. This comprehensive approach aimed to enhance the performance and robustness of the models in both classification and regression tasks.

#### - Classification Models

For the classification task, three ML algorithms are employed: Linear logistic regression classifier (LogReg) (Hastie et al., 2009), random forest (RF) classifier (Breiman, 2001), and support vector machine (SVM) (Cortes & Vapnik, 1995). LogReg models are designed for predicting probabilities of *K* classes or categories in the classification problem via linear functions of input features *x*, ensuring they sum to one and stay within a valid probability range of [0, 1]. The model is expressed in terms of *K* − 1 log-odds or logit transformations and uses the last class as the denominator in the odds-ratios, with a special simplicity in binary classification scenarios (Hastie et al., 2009). RF is an ensemble learning method that builds multiple decision trees by randomly sampling with replacement from the training data. Each tree is grown by recursively selecting random feature subsets and identifying optimal split-points. For classification, the final output is determined by the majority vote of individual tree predictions during prediction. Thus, the model output is an ensemble of trees that collectively make predictions. Hyperparameters include the number of trees in the forest, the maximum depth of the trees, and the minimum number of samples required to split a node (Hastie et al., 2009). The SVM algorithm classifies data by transforming each data point into a k-dimensional feature space, where k >> *n* and *n* is the number of features. It identifies a hyperplane that maximizes the margin between classes, minimizing classification errors. The margin is the distance between the decision hyperplane and the nearest instance of each class (Uddin et al., 2019).

These algorithms were chosen due to their well-established competitive performance in classification tasks and adaptability to diverse scenarios. For instance, Uddin et al. (2019) provide a comprehensive overview of the relative performance of different supervised ML algorithms for disease prediction, including LogReg, RF, and SVM. Their findings, despite variations in frequency and performance, underscore the potential of these algorithmic families in disease prediction.

Certain model hyperparameters were fixed for each algorithm while others were optimized using cross-validation. For the **LogReg** algorithm, an intercept term, which represents the log-odds of the baseline class, was included in the model by the default setting of scikit-learn. The parameter class_weight was set as balanced to adjust the weights assigned to classes during the training process. This indicates that the algorithm automatically adjusts the weights of the classes inversely proportional to their frequencies. The maximum number of iterations taken for the solver to converge was set to 500. Solver here refers to the optimization algorithm used to find the optimal values for the coefficients of the linear logistic regression model. L2-norm regularization was applied to prevent overfitting and improve the generalization of the model. Regularization involves adding a penalty term to the loss function, and in L2-norm regularization, the penalty is proportional to the square of the magnitudes of the coefficients. The regularization parameter C was set to values 0.0001, 0.001, 0.01, 0.1, 1, 10, and 1000. The optimal hyperparameter as identified using cross-validation is C = 0.1.

For the **RF classifier**, model hyperparameters include the criterion used by each decision tree to determine where to split and the maximum number of features to consider for the best split, set to the square root of the total number of features. Specifically, the criterion hyperparameter was configured to use the Gini diversity index. The Gini index is employed by each decision tree in the RF ensemble as the measure for determining optimal split points during the training process. For tree building, a bootstrap method was used, involving sampling with replacement instead of using the whole training set to build trees. No constraints were imposed on the depth of the trees. The hyperparameter tuning of the number of trees (n_estimators) was set to values 100, 500, 1000, 5000, and 10000. The optimal model’s n_estimators as identified using cross-validation is 500.

For the **SVM** algorithm, the parameter class_weight was set as balanced. Two variants of the SVM algorithm were tested. The first one used a non-linear radial basis function (rbf) kernel with kernel width gamma (kernel: rbf). Tested hyperparameter values included: C: 0.01, 0.1, 1, 10, 100, 500, 1000; gamma: 0.00001, 0.0001, 0.001, 0.01, 0.1, 1, 10. The second variant used a parameterless linear kernel. Tested hyperparameter values for this choice included: C: 0.01, 0.1, 1, 10, 100, 500, 1000; kernel: linear. The optimal combination of hyperparameters as identified using cross-validation are C: 1000, gamma: 1e-05, kernel: rbf.

Results of the hyperparameter tuning and grid search for each classifier can be found in the Supplementary Material (**Tables S1-S3**, **Figures S2-S4**).

Benchmarking was conducted using dummy models representing baseline references. Baseline models serve as the null hypothesis, simulating scenarios where models possess minimal knowledge about individual sample labels, essentially reflecting the label distribution in the training data at best. These models are instrumental for assessing whether a sophisticated model’s performance surpasses the “null” or “chance” level, providing a benchmark to evaluate the effectiveness of advanced algorithms.

Baseline models consisted of a uniform random baseline (randomly assigning class labels with equal probability, i.e., 50% for each class, without considering any input features), a most frequent baseline (it assigns the majority class label to all instances), and a stratified random baseline (which randomly assigns labels proportionally to their relative frequency in the training data) (Murphy, 2012).

#### - Regression Models

For the regression task, three ML algorithms are utilized: Ridge regression (RR) (Hastie et al., 2009), RF regressor (Breiman, 2001), and support vector regressor (SVR) (Cortes & Vapnik, 1995). RR, often referred to as L2-norm regularized least-squares regression, adds a regularization term to the standard linear regression objective function. In RR, the objective function to be minimized is the sum of the squared differences between the observed and predicted values (least squares term), and a penalty term that discourages overly complex models by adding the L2-norm of the coefficients multiplied by a regularization parameter (alpha). The regularization term penalizes large coefficients, preventing overfitting and promoting a more stable and generalizable model. The strength of this penalty is controlled by the hyperparameter alpha (Hastie et al., 2009). RF constructs multiple decision trees by bootstrapping the data and introducing randomness in variable selection at each node. The final prediction is an aggregation of predictions from all individual trees. For regression, the final output is determined by the average of individual tree predictions during prediction (Hastie et al., 2009). SVR uses the principles of SVM for regression tasks. Similar to the latter, SVR introduces the concept of a margin. In SVR, the margin represents a range of values within which errors are tolerable. SVR aims to fit the function *f*(*x*) in such a way that the differences between the predicted values and the actual values (errors) fall within the specified margin. SVR minimizes the errors while staying within the margin. The loss function penalizes deviations from the actual values but allows for a certain amount of error within the margin. To prevent overfitting, SVR includes a regularization term controlled by the parameter C, which is a user-defined hyperparameter. The regularization term controls the smoothness of the learned function (Hastie et al., 2009).

Certain model hyperparameters were fixed for each algorithm while others were optimized using cross-validation. For the **RR** algorithm the regularization strength parameter, denoted as alpha, was set to values 0.0001, 0.001, 0.01, 0.1, 1, 10, 100, and 1000. The optimal model’s alpha as identified using cross-validation is 100.

For the **RF regressor**, model parameters include the criterion of mean squared error (measuring the quality of estimators’ split), and the maximum number of features considered for the best split, which is set to 1, as empirically justified in Geurts et al. (2006). For tree building, a bootstrap method was used, involving sampling with replacement instead of using the whole training set to build trees. No restrictions were imposed on the depth of the trees. The hyperparameter tuning of the n_estimators was set to values of 100, 500, 1000, 5000, and 10000. The optimal model’s n_estimators parameter as identified using cross-validation is 5000.

For the **SVR** training, the shrinking trick described in Chang and Lin (2011) was employed. Shrinking attempts to reduce the optimization problem by removing the elements that have already been bound. Two variants of the SVR algorithm were tested. The first one used a rbf kernel and the hyperparameters C, epsilon, and gamma are varied. The tested values included: C: 0.01, 0.1, 1, 10, 100, 500, 1000; epsilon: 0.001, 0.01, 0.1, 1; gamma: 0.00001, 0.0001, 0.001, 0.01, 0.1, 1, 10; kernel: rbf. The second variant used a linear kernel and only the hyperparameters C and epsilon are varied. The hyperparameters values for this choice included: C: 0.01, 0.1, 1, 10, 100, 500, 1000; epsilon: 0.001, 0.01, 0.1, 1; kernel: linear. The optimal hyperparameters combination as identified using cross-validation are C: 500, epsilon: 0.1, gamma: 0.00001, kernel: rbf.

Results of the hyperparameter tuning and grid search for each regressor can be found in the Supplementary Materials (**Tables S4-S5**, **Figures S5-S7**).

Similar to the classification models, dummy models were included as benchmark references for regression tasks. These models encompassed a randomly shuffled baseline (predicting the target variable by randomly shuffling actual target values), a mean and a median baseline (predicting the arithmetic average and the median value of the target variable, respectively, for all instances, without considering any input features or patterns) (Murphy, 2012).

#### - Evaluation of Classification Model Performance

The performance of the classification models on test data was assessed using five metrics: Area Under the Precision-Recall Curve (AUPRC), Area Under the Receiver Operating Characteristic curve (AUROC), Balanced Accuracy, F1 score, and Brier loss. These metrics all range from 0 to 1, where 0 indicates worst and 1 indicates best performance. An exception is Brier loss, for which 0 indicates perfect model performance and uncertainty calibration, while a loss of 1 indicates the worst performance.

Precision measures a classifier’s ability to avoid incorrectly labeling negative examples as positives, while Recall (also known as “sensitivity” or “true positive rate”) measures its ability to identify all positive examples correctly as positives (Powers, 2011). The AUPRC represents the tradeoff between Precision and Recall over all possible classifier thresholds as quantified by the area under the Precision curve integrated over all Recall values. The AUROC represents the tradeoff between Recall and the true negative rate (also known as “specificity”, Davis & Goadrich, 2006; Fawcett, 2006). The Balanced Accuracy score calculates the average of sensitivity (true positive rate) and specificity and it is used to ensure that the model’s performance on imbalanced datasets is not overestimated. The F1 score is the harmonic mean of Precision and Recall scores (Powers, 2011). The Brier loss is calculated as the mean squared difference between the predicted probability and the true binary label (Brier, 1950). It measures whether the probabilities provided by the model are well-calibrated. These predicted probabilities are derived from the model’s internal mechanisms, which assess the input features and generate probability estimates for each instance in the dataset.

#### - Evaluation of Regression Model Performance

To assess the performance of the regression models on test data, three key metrics were employed: R2, Mean Absolute Error (MAE), and Mean Squared Error (MSE). The R2 score measures the proportion of variance in the target variable explained by the model and is calculated as 1 minus the ratio of the model’s MSE to that of a mean baseline model. The R2 score is a coefficient of determination and, theoretically, it can range from -∞ to 1, where 1 indicates perfect prediction, 0 indicates no improvement over the mean model, and values less than 0 indicate poorer performance than the baseline (Bishop, 2006; James et al., 2017). MSE and MAE represent the average squared and absolute differences between predicted and actual scores, respectively, and are used to assess the model’s prediction accuracy (Bishop, 2006; James et al., 2017). For both MSE and MAE, lower values signify better model performance.

#### - Bootstrapping with Replacement

To enhance the reliability of performance evaluation, bootstrapping with replacement was utilized. Specifically, we resampled the test set 100 times with replacement to create distributions of performance estimates. Each resampled sample had the same dimensions as the original sample, and mean performance estimates with 95% confidence intervals (CI) were derived using this method (Davison & Hinkley, 1997).

#### **-** Feature Importance Analysis: Comparing Multi- vs. Univariate Models for Depression Prediction

Feature importance analysis was conducted to assess the predictive strength of each individual feature with respect to the target variable, depression. For this analysis, all ML algorithms previously employed for the classification (LogReg, RF, and SVM classifiers) and regression (RR, RF regressor, and SVR) tasks were applied to single features. Using different algorithms can provide a more comprehensive view of feature importance. The purpose of this analysis was to determine the extent to which individual features are predictive of depression.

Here, the performance of multivariate models, which incorporate all features simultaneously, is compared with that of univariate models, each including a single feature. The metrics used for this comparative analysis are the mean AUPRC score (with 95% CI) for classifiers and the mean R2 score (with 95% CI) for regressors. In this assessment, each multivariate model, whether for classification or regression tasks, serves as a benchmark. This enables an evaluation of whether the collective impact of features enhances prediction accuracy.

### Ethical Considerations

This study was approved by the Ethics Committee of the Psychological Research Institute, Universidad Nacional de Córdoba (CEIIPsi-UNC-CONICET), 14/02/20–23/03/20. The privacy and confidentiality of participants’ data were ensured, and informed consent was obtained from all participants before their participation. Guidelines and regulations regarding the use of human subjects in research were also followed.

## RESULTS

### 1. Classification Models: Identifying Depression Presence

The performance of ML classifiers for predicting depression in college students during the COVID-19 pandemic is evaluated across five metrics and compared to the metrics of dummy baseline models. The baseline models serve as sanity checks for evaluating the performance of the ML classifiers.

Figure 1 summarizes the performance metrics for both the trained and tested ML models and the tested baseline models. As expected and across all metrics, the ML models during the test phase achieve a superior predictive ability using the provided input features compared to random chance.

**Figure 1.**
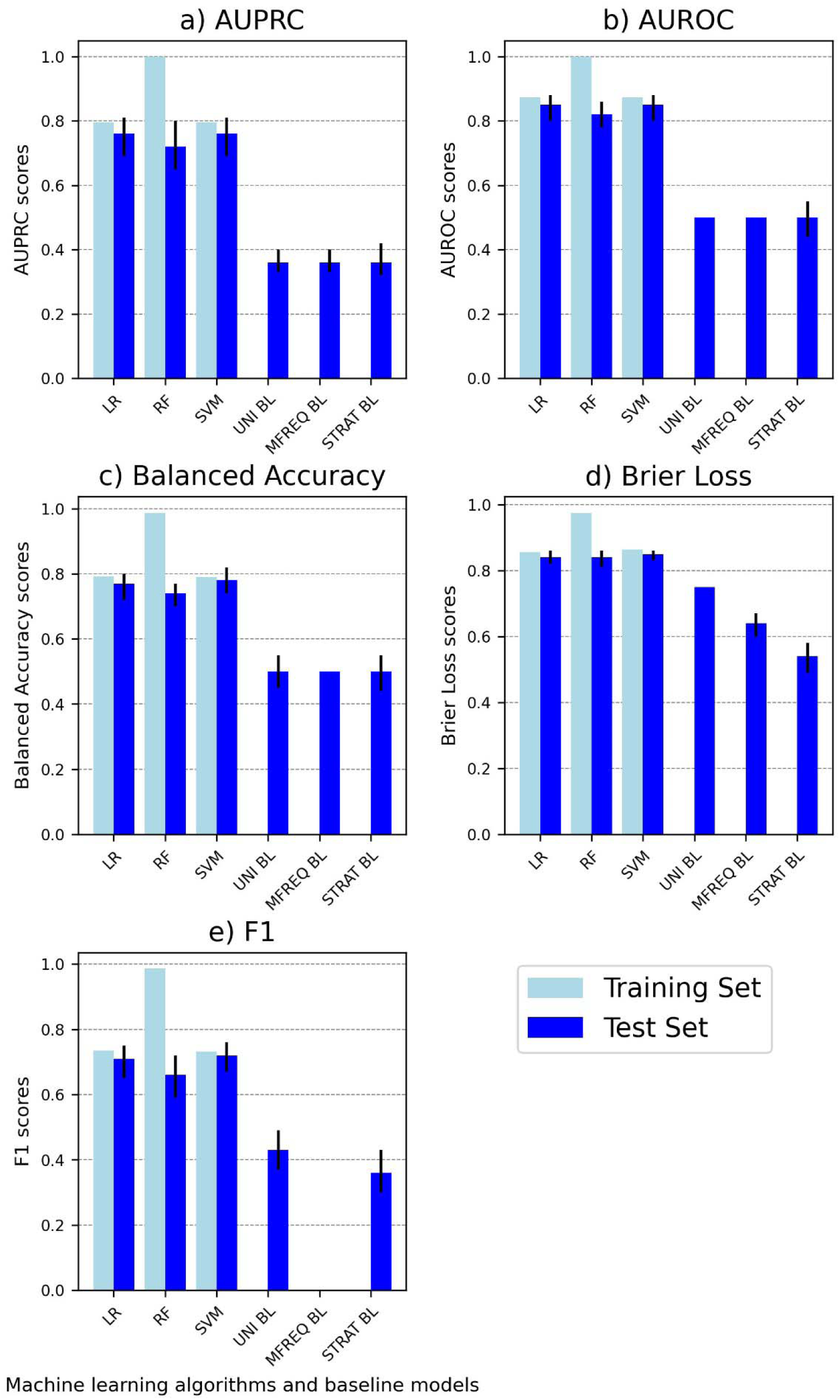
Performance of machine learning classifiers on the training and test sets as well as of baseline classifiers on the test set. The bar charts depict the performance of machine learning classifiers (linear logistic regression, random forest classifier, and support vector machine) on the training set and the test set and of baselines classifiers (uniform random, most frequent, and stratified random) on the test set using five performance metrics: **(A)** AUPRC, **(B)** AUROC, **(C)** Balanced accuracy, **(D)** Brier loss, and **(E)** F1. Light blue bars represent performance scores on the training set, while blue bars with error bars represent mean test set scores with 95% confidence intervals. To ensure that higher values consistently indicate better performance across all methods, the Brier loss is plotted as 1-Brier. *Abbreviations:* AUPRC: Area Under the Precision-Recall Curve. AUROC: Area Under the Receiver Operating Characteristic Curve. LR: Linear logistic regression. RF: Random forest classifier. SVM: Support vector machine. UNI BL: Uniform random baseline. MFREQ BL: Most frequent baseline. STRAT BL: Stratified random baseline.

The SVM classifier is on par with or outperforms both alternative classifiers, attaining the highest AUPRC score of 0.76 (95% CI: 0.69, 0.81) and the lowest Brier score loss of 0.15 (0.14, 0.17). The LogReg classifier closely follows, displaying an identical AUPRC score of 0.76 (0.69, 0.81) and a slightly higher Brier score loss of 0.16 (0.14, 0.18).

The RF classifier generally achieves lower performance than SVM and LogReg, attaining AUPRC score of 0.72 (0.65, 0.80) and Brier score loss of 0.16 (0.14, 0.19). RF also shows evidence of overfitting as indicated by a rather substantial drop in AUPRC performance from 1 (0.9995) on the training set to 0.72 on the test set. Refer to Figure 1 and **Table S6** for a visualization of these and the remaining performance metrics (i.e., AUROC, Balanced accuracy, and F1 scores) employed in this study for each ML classifier.

Refer to Figure 2 for the Precision-Recall Curves and Receiver-Operating Characteristic depicting depression prediction. This plot aids in comparing models’ ability to distinguish between positive and negative classes (Depressed and Non-Depressed, respectively) at different clinically relevant operating points such as high sensitivity and high specificity). Confusion matrices for each ML classifier are displayed in the Supplementary File (**Figure S8**).

**Figure 2.**
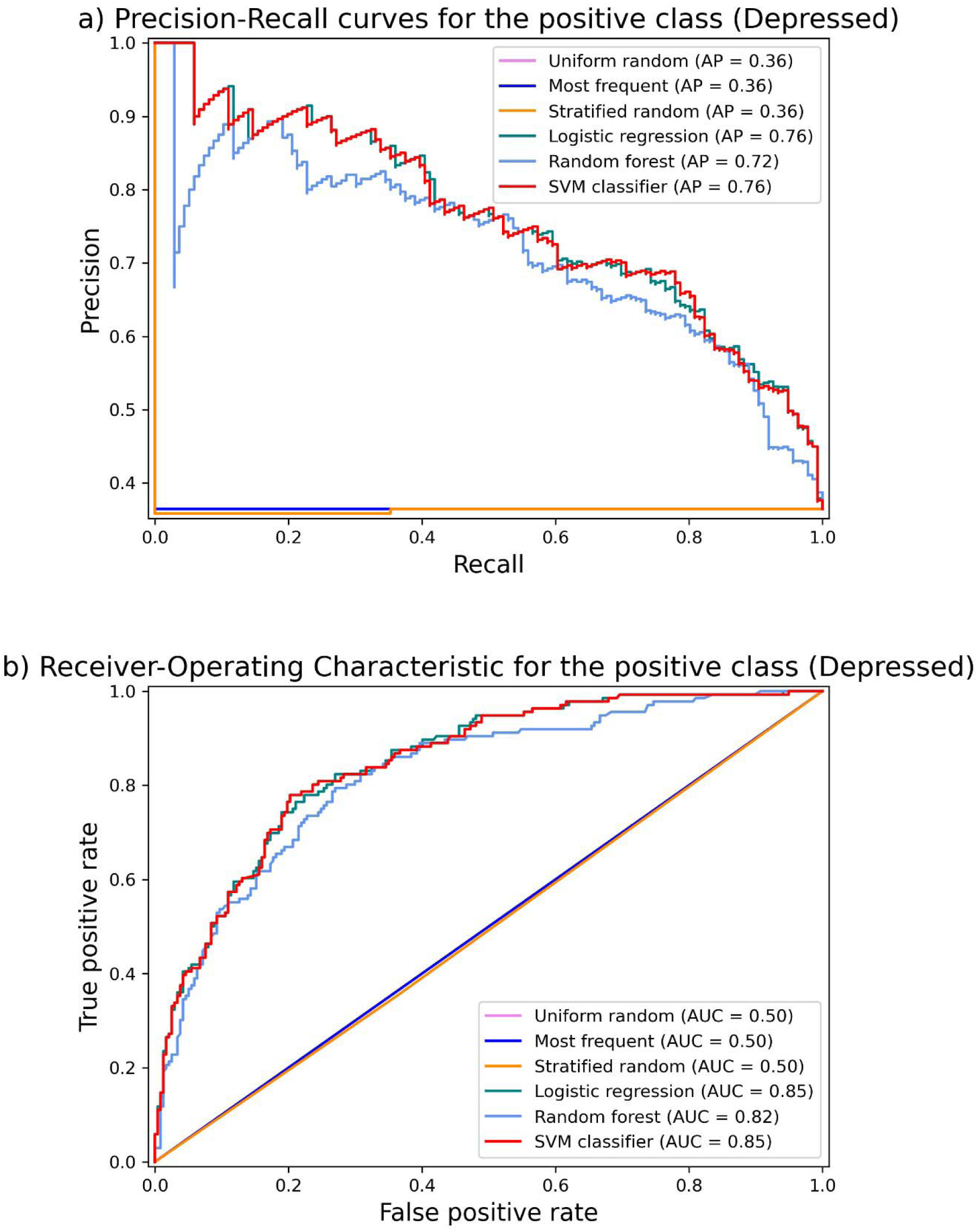
Classifier performance comparison: Precision-Recall Curves and Receiver-Operating Characteristic Curves for depression prediction. These curves characterize the predictive performance of trained models (linear logistic regression, random forest classifier, and support vector machine) and dummy classifiers (uniform random, most frequent, and stratified random baselines) for depression prediction for varying classifier thresholds, where the positive class indicates depression presence. (A) Precision-Recall Curve (PRC) displaying how precision changes with different recall levels for various models/baselines. (B) Receiver-Operating Characteristic (ROC) curves illustrating the balance between true positive rate (sensitivity) and false positive rate (1-specificity) as probability thresholds change. Each curve depicts how sensitivity changes with different specificity levels for different models/baselines. *Abbreviations*: SVM: Support vector machine. AP: Precision-Recall curve values (the Precision-Recall curve display in scikit-learn uses the term “average precision” (AP) to refer to the area under the precision-recall curve). AUC: Area Under the Receiver Operating Characteristic values.

### 2. Regression Models: Quantifying Depression Severity

The performance of ML regressors in predicting depression among college students during the COVID-19 pandemic is evaluated across three key metrics and compared to the results of these metrics on dummy baseline models. The baseline models serve as benchmarks for evaluating the performance of the ML regressors.

Figure 3 provides an overview of the performance metrics for both the trained and tested ML models, along with the tested baseline models. As expected, the tested ML models consistently demonstrate superior predictive capabilities using the provided input features compared to random chance, as evidenced by all metrics.

**Figure 3.**
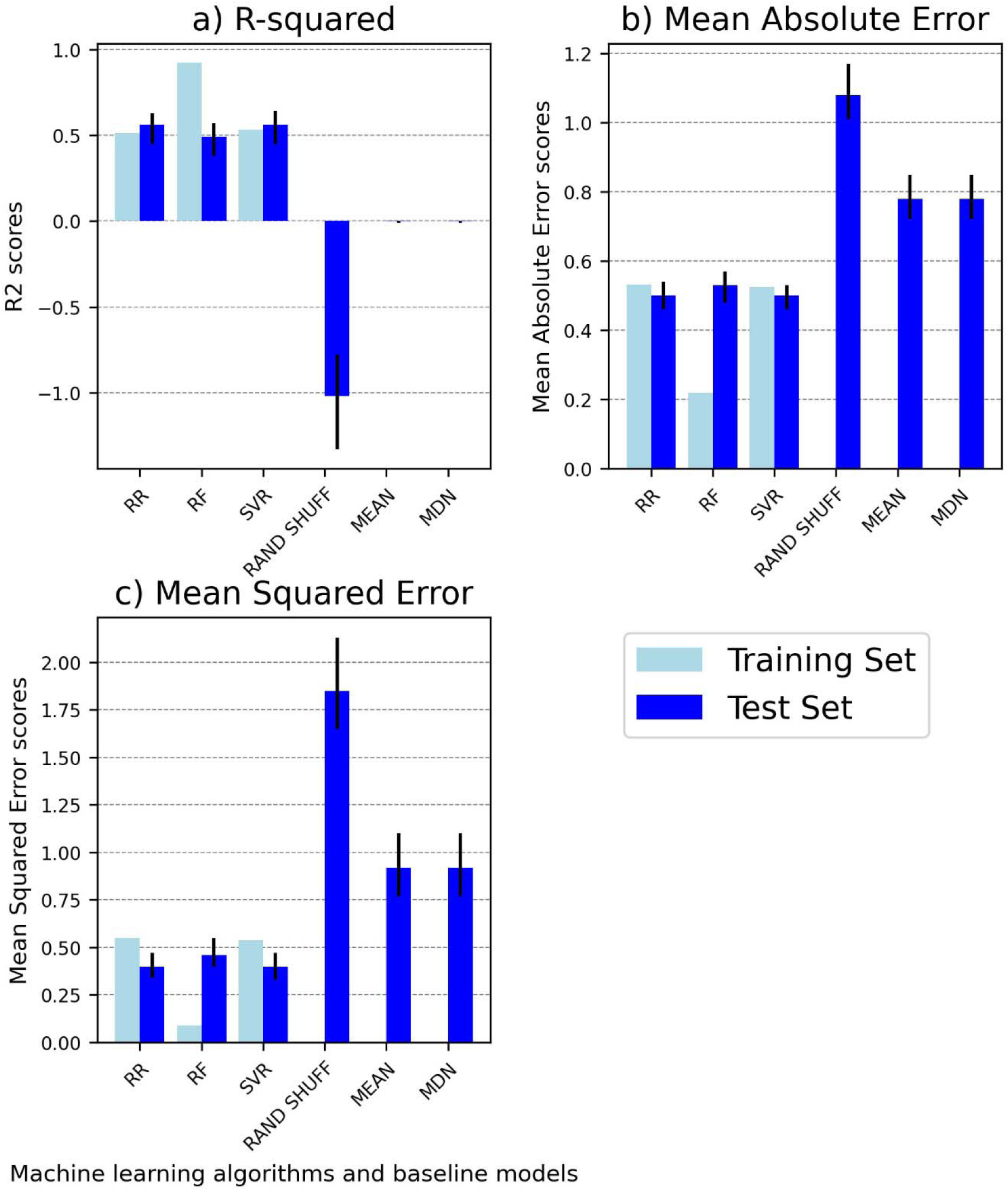
Performance comparison of machine learning regressors on the training and test sets and baselines on the test set. The bar charts compare the performance of machine learning regressors (ridge regression, random forest regressor, and support vector regressor) on the training set and the test set and of baseline regressors (randomly shuffled baseline, mean baseline, and median baseline) on the test set using three performance metrics: **(A)** R-squared, **(B)** Mean Absolute Error, **(C)** Mean Squared Error. Light blue bars represent performance scores on the training set, while blue bars with error bars represent mean test set scores with 95% confidence intervals. *Abbreviations:* R2: R-squared. RR: Ridge Regression. RF: Random Forest Regressor. SVR: Support Vector Regressor. RAND SHUFF: Randomly Shuffled Baseline. MEAN: Mean Baseline. MDN: Median Baseline.

SVR and RR achieve the highest R2 score of 0.56 (95% CI: SVR: 0.45, 0.64 and RR: 0.45, 0.63). This R2 value indicates that the models can account for approximately 56% of the variability in the target variable. The RF regressor achieves a lower R2 value of 0.49 (0.38, 0.57).

The RF regressor also exhibits inferior MAE and MSE performance on the test set compared to the SVR and RR models. A comparison of the RF regressor’s performance between the training and test sets reveals a high degree of overfitting, indicating that the complexity of the model was too high to ensure robust generalization. Both the MAE and MSE metrics for the RR algorithm mirror the SVR’s performance, underscoring their comparable predictive capabilities (Figure 3; **Table S7**). The correlations between actual and predicted values for each ML algorithm in the test set are depicted in the Supplementary File (**Figure S9**).

### 3. Evaluation of Feature Importance: Comparison of Multivariate to Univariate Models

#### 3.a. Comparison of Multivariate to Univariate Models in the Classification

##### Task

In the classification task for predicting depression presence, the multivariate model generally outperforms univariate models, except for the RF classifier using depression scores (at T1) as a single feature (Figure 4, **Table S8**). However, it’s crucial to note that the multivariate model for the RF classifier shows signs of overfitting (Figure 1).

**Figure 4.**
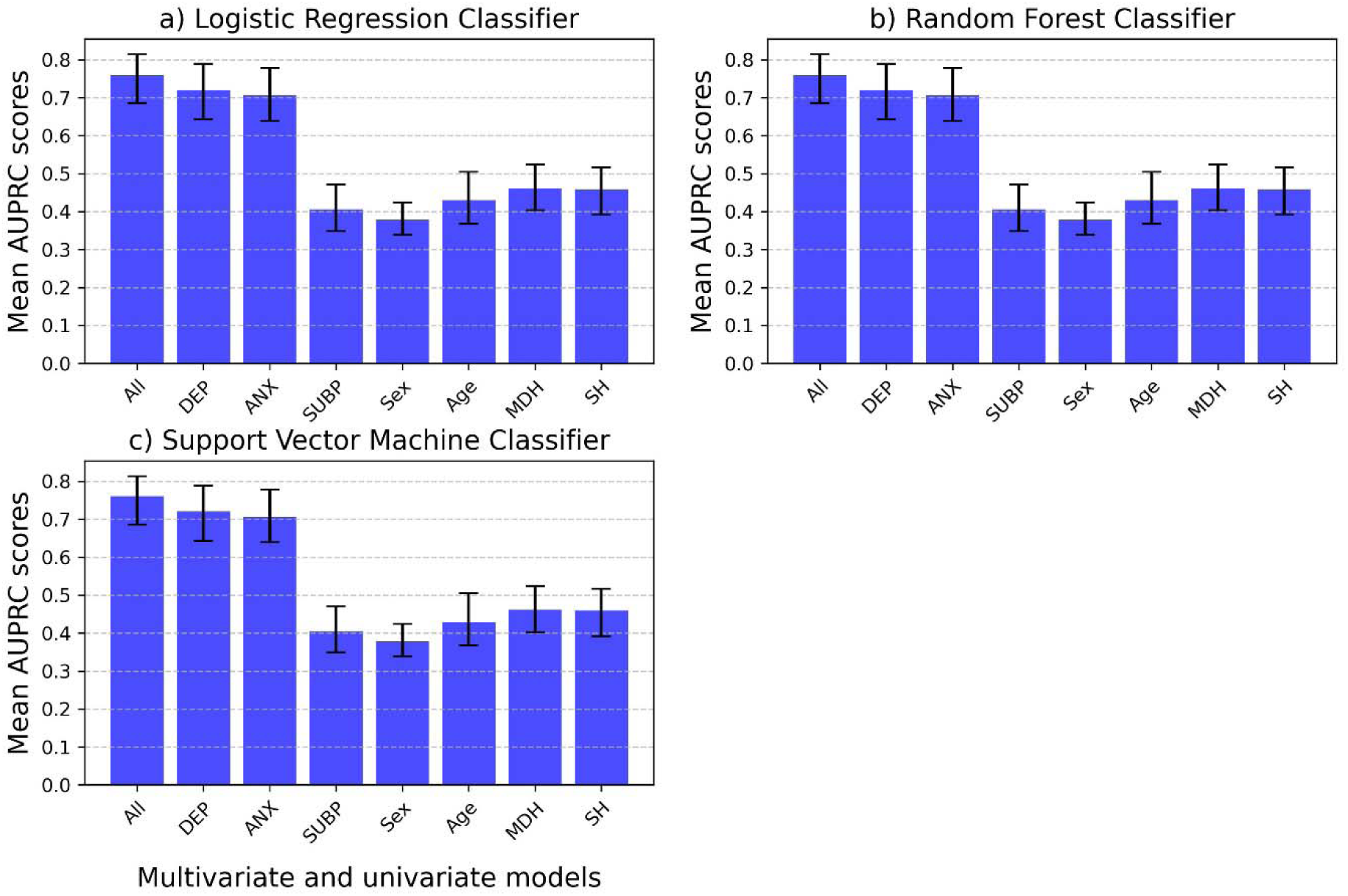
Mean AUPRC scores for multivariate versus univariate machine learning models predicting depression in the classification task. Linear logistic regression, random forest and support vector machine classifiers were trained either as multivariate models (encompassing all features) or univariate models (each incorporating just a single feature). Error bars represent 95% confidence intervals. *Abbreviations*: AUPRC: Average Precision-Recall Curve; All: Multivariate model with all features included; Univariate models: DEP: Depression scores measured at time 1 as the single feature; ANX: Anxiety scores as the single feature; SUBP: Quarantine sub-periods as the single feature; Sex: Biological sex as the single feature; Age: Age as the single feature; MDH: Mental disorder history as the single feature; SH: Suicidal behavior history as the single feature.

Univariate classifiers, in particular LogReg and SVM using depression (at T1) or anxiety scores as single features, exhibit comparative performance levels (in terms of mean test AUPRC) that are close to those of the corresponding multivariate model that include all features. On the other hand, all other univariate models perform significantly worse, with mean AUPRC scores slightly above (models using age, mental disorder history, and suicidal behavior history as single features) or approaching chance-level performance (models using quarantine sub-periods and sex as single features). Importantly, this trend is consistent across the three ML algorithms tested (Figure 4, **Table S8**).

#### 3.b. Comparison of Multivariate to Univariate Models in the Regression Task

In the regression task for predicting depression scores, the multivariate model consistently outperforms all univariate models. Notably, when considering all univariate models, depression (at T1) and anxiety scores emerge as the most predictive single features, achieving the highest performance levels (mean R2 scores), particularly when using the RR algorithm. It is noteworthy that univariate models excluding both depression (at T1) and anxiety scores exhibit performance levels slightly above (e.g., suicidal behavior history) or comparable (e.g., quarantine sub-period) to chance-level. Importantly, this pattern is consistent across the three ML algorithms. Contrary to the results on the comparison between multivariate classifiers performance, when comparing univariate models performance, RF models perform better than SVR models. (Figure 5, **Table S9**).

**Figure 5.**
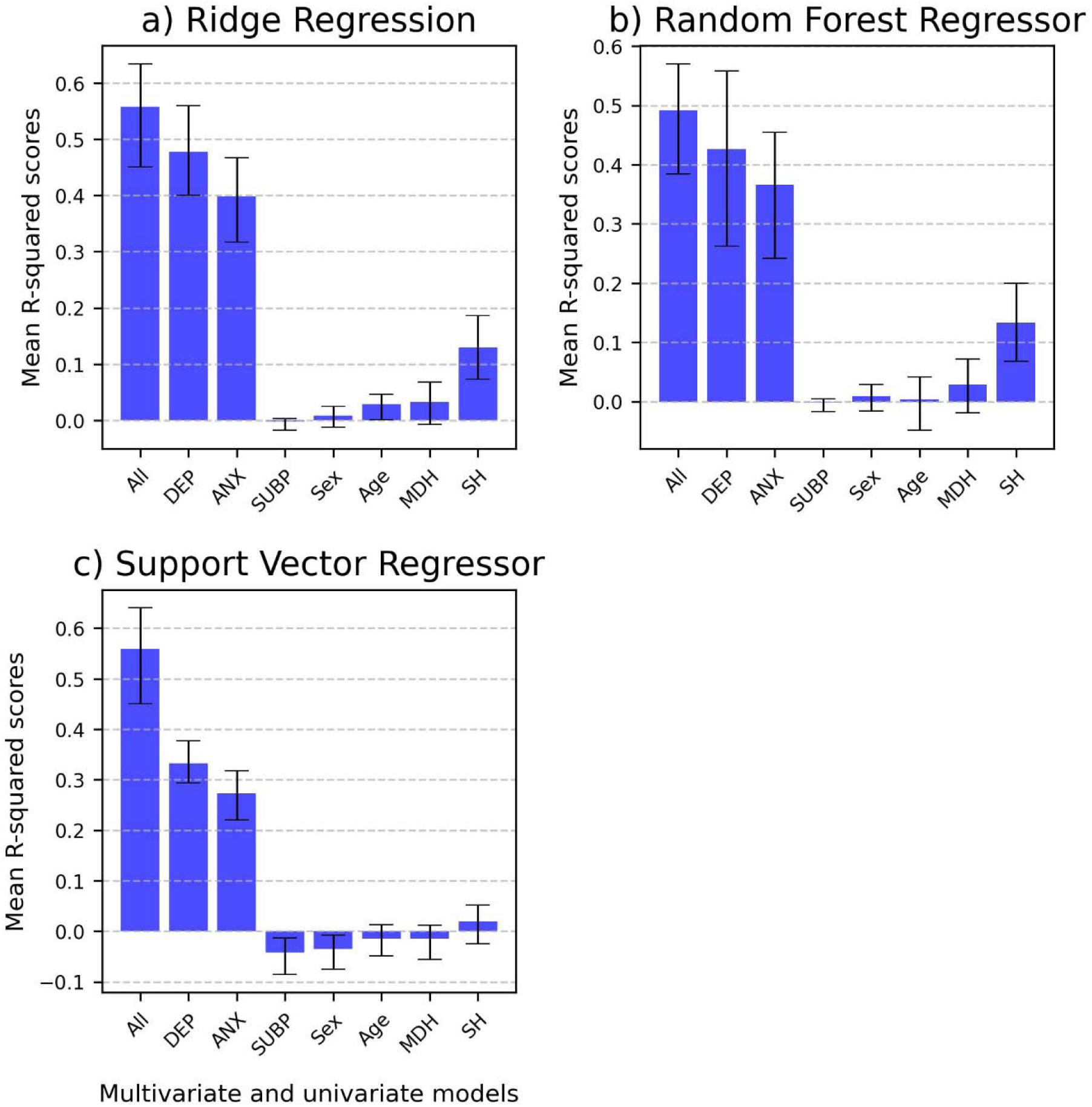
Comparative analysis of mean R-squared scores for multivariate versus univariate machine learning models predicting depression in the regression task. Bar plot illustrating the comparison of mean R2 scores for various machine learning models (ordinary least-squares regression, random forest regressor, and support vector regressor) in the regression task of predicting depression. The models evaluated include multivariate models (encompassing all features) and univariate models (each incorporating a single feature). The bar plots include error bars representing 95% confidence intervals and using markers to highlight data points. *Abbreviations*: All: Multivariate model with all features included; Univariate models: DEP: Depression scores measured at time 1 as the single feature; ANX: Anxiety scores as the single feature; SUBP: Quarantine sub-periods as the single feature; Sex: Biological sex as the single feature; Age: Age as the single feature; MDH: Mental disorder history as the single feature; SH: Suicidal behavior history as the single feature.

## DISCUSSION

This study leveraged a longitudinal dataset for the prediction of depression among college students amidst the challenging backdrop of the COVID-19 pandemic, allowing for the examination of evolving mental health patterns over time. The results indicate a gain in predictive efficacy of multivariate ML algorithms as compared to their univariate counterparts.

As previously mentioned, the dataset analyzed using ML in this study underwent scrutiny through MEM in a prior study (López Steinmetz et al., 2023). In that study, variables such as sex (female), age (younger), mental disorder history (presence), and suicidal behavior history (presence) were identified as having significant main effects on depression. However, following Cohen’s conventions for effect sizes (ES; Cohen, 1988), these effects were deemed small for sex (ES = 0.09), age (ES = 0.09), and mental disorder history (ES = 0.17), while being of medium magnitude for suicidal behavior history (ES = 0.34) (López Steinmetz et al., 2023). Notwithstanding, the outcomes of that prior MEM study align with existing literature, underscoring that variables associated to adverse mental health effects after and during the COVID-19 pandemic encompass baseline depression, female sex, younger age, the presence of mental disorders, and student status, among others (Liu et al, 2019; Xiong et al., 2020). In the current study, the application of ML models enabled the exploration of various features’ predictive potential, aligning with those examined in the previous study (López Steinmetz et al., 2023). Notably, anxiety, a feature included in this ML-based approach but absent in the previous study, revealed as one of the most relevant features, alongside depression at T1, for identifying depression presence and quantifying depression severity. The significance of anxiety as a predictor of depression underscores the interconnectedness of these mental health conditions. It aligns with existing literature that emphasizes the comorbidity and shared features between anxiety and depression (Brown et al., 2001; Kalin, 2020; Lamers et al., 2011) particularly in college students during COVID-19 (Bai et al., 2021; Chang et al., 2021). As for depression at T1, these findings suggest that depressive symptoms persist over time during the COVID-19. This aligns with research conducted in college students (Zhao et al., 2023) and general populations in developed countries (Ettman et al., 2022) and Argentina (Canet-Juric et al., 2020), demonstrating the enduring nature of depression symptoms amid the pandemic.

As for the ML algorithms performance in this study, both SVM/SVR and LogReg/RR demonstrated the best results in both classification and regression tasks. SVM has consistently been identified as a high-performing algorithm in predicting depression among students (see, e.g., Choudhury et al., 2019; Gil et al., 2022; Qasrawi et al., 2022). However, RF also emerged as the top-performing algorithm in some studies (see, e.g., Gil et al., 2022; Qasrawi et al. 2022; Rois et al., 2021), which contrasts with the present findings where it displayed overfitting and exhibited comparative lower performance. The observed overfitting in the RF algorithm in both classification and regression tasks within this study can be attributed to various factors. Although RFs are renowned for their adaptability and capacity to discern intricate data relationships, this characteristic may lead to overfitting, particularly if the model’s complexity surpasses the dataset’s requirements. The profusion of decision trees and their interactions may contribute to an overfitted model. Additionally, the hyperparameters of the RF, such as the number of trees, maximum depth of trees, and other parameters, play pivotal roles. In this study, only a range of values for the number of trees were explored. However, it’s possible that other hyperparameters, like the maximum depth of the trees, might need further exploration to find the right balance between model complexity and generalization. Likewise, RFs automatically perform feature selection by considering a subset of features at each split. While advantageous, this process can lead to overfitting if the model emphasizes noise or irrelevant features. In this study, an examination of multi-versus univariate models revealed two dominant features (depression at T1 and anxiety), with the remaining features exhibiting limited significance. As for the discrepancies in algorithm performance between different studies, these may be ascribed to several factors. To mention some of them, the diverse array of features integrated into the models can significantly impact performance when comparing studies. Additionally, variations in study designs, predominantly the widespread use of a cross-sectional design in studies analyzing questionnaire-collected data, further contribute to these differences. Depression assessment methods, relying on self-reported and clinician-administered questionnaires, exhibit inherent limitations (Fried & Nesse, 2015). However, it is crucial to emphasize that, particularly in developing countries, accessing potent features capable of identifying biomarkers, such as those related to neuroimaging (Wise et al., 2014), is often impeded by their high cost. Consequently, there is a pressing need to identify reliable and cost-effective predictors to develop risk prediction models capable of enhancing clinical decision-making in depression diagnosis and care.

In addition, addressing the challenge of interpretability in ML models is crucial for their effective deployment. This challenge is particularly pronounced in the transparency of decision-making processes within advanced models, often referred to as “black-boxes” (Ribeiro et al., 2016). Notably, complex models like RF, which leverage numerous decision trees in a non-linear interaction, exacerbate the difficulty in comprehending underlying mechanisms (Breiman, 2001). In contrast, linear models such as logistic regression and RR are frequently considered to be easier to interpret due to their transparent mathematical formulations (Hosmer et al., 2013). Nevertheless, even the interpretation of multivariate linear models and decision trees can be highly misleading (Haufe et al., 2014; Wilming et al., 2022; Wilming et al., 2023). Moreover, additional challenges persist in high-dimensional datasets (Belloni et al., 2014). For these reasons, we here resort to univariate analyses to unanimously clarify the aptitude of individual features for prediction (Haufe et al., 2014). In this study, while multivariate ML models encompassing all features demonstrate robust performance in both classification and regression tasks, particularly with LogReg/RR and SVM/SVR, a closer examination reveals a reliance on two key features: depression (at T1) and anxiety. The relevance of these variables in identifying depression aligns with existing literature (American Psychiatric Association, 2000; Cassady et al., 2019; Horn & Wuyek, 2010; Liu et al, 2019).

However, this study’s outcomes also emphasize the limited significance of certain variables, encompassing quarantine duration and restrictiveness (referred to as quarantine sub-periods), sex, age, mental disorder history, and suicidal behavior history, not only in identifying depression presence (i.e., classification task), but particularly in quantifying depression severity (i.e., regression task). This departure from established psychological literature, which traditionally links these variables to depression prediction and diagnosis both before and during pandemics (e.g., Kisch et al., 2005; Nolen-Hoeksema & Aldao, 2011; Piccinelli & Wilkinson, 2000; Sheldon et al., 2021; Solomou & Constantinidou, 2020; Brooks et al., 2020), is noteworthy. Nevertheless, it is also worth noting that some recent research, such as a systematic review and meta-analysis, found no significant association between sex and depression among undergraduates (Yang et al., 2023). Overall, the presented findings suggest that, when considered in isolation, these variables may not reliably predict depression within this population. This underscores the need for a more holistic approach to depression prediction, emphasizing the intricate interplay of multiple factors.

The observed disparities among these current ML-based results, prior MEM analysis, and existing literature bear implications for translational research, as well as for health decision-makers and policymakers striving to improve mental health, particularly to offer scalable and effective evidence-based interventions addressing depression (Meehan et al., 2022). Reliable predictors and prediction models are crucial for effective mental health interventions, particularly those targeting depression prevention (Bernardini et al., 2017). While internal validation provides strong support for the performance of the models tested in this study, it is imperative to underscore the necessity for additional external validation and replication studies to solidify the implications of these findings. The prevailing landscape of developing clinical prediction models related to mental disorders calls for more rigorous validation procedures. The current studies in this field often exhibit a deficiency in both external and internal validation (Meehan et al., 2022), emphasizing a critical weakness that needs to be addressed in future research.

Especially in the post-COVID-19 era, there is a growing imperative to leverage advanced technologies like ML algorithms and artificial intelligence to enhance the precision of depression screening among college students. This, in turn, can enable proactive prevention efforts and contribute significantly to improving mental health outcomes (Liu et al., 2022).

In conclusion, this study not only advances our understanding of depression prediction in college students from a developing country using ML algorithms, but also highlights the need for further exploration and validation of predictive models in diverse populations. Future research should aim to refine models, consider additional psychological and socio-environmental factors, and enhance external validation to ensure the broad applicability of these findings. This way, the implementation of scalable and effective interventions for diagnosing and preventing depression may become more attainable.

## Disclosure statement

No potential conflict of interest was reported by the authors.

## Data availability statement

The reproducible Python code is available in the Open Science Framework (OSF) repository, https://doi.org/10.17605/OSF.IO/QT2GU, and in the GitHub repository, https://github.com/cecilost/machine-learning-to-predict-depression-in-college-students-during-the-COVID-19-pandemic. The dataset is available in the OSF repository, https://doi.org/10.17605/OSF.IO/2V84N.

## Funding

This result is part of a project that has received funding from the European Research Council (ERC) under the European Union’s Horizon 2020 research and innovation programme (Grant agreement No. 758985).

## Notes

### Competing Interest Statement

The authors have declared no competing interest.

### Author Declarations

This study was approved by the Ethics Committee of the Psychological Research Institute, Universidad Nacional de Cordoba, 14/02/20 23/03/20. The privacy and confidentiality of participants' data were ensured, and informed consent was obtained from all participants before their participation. Guidelines and regulations regarding the use of human subjects in research were also followed.

