## Supplemental File for "Machine Learning Models Predict the Emergence of Depression in Argentinean College Students during Periods of COVID-19 Quarantine"

**INDEX**

| **Figure S1.** Description of the sampling design. | 3 |
| --- | --- |
| **Table S1.** Results of the hyperparameter tuning for the Linear Logistic Regression Classifier | 4 |
| **Figure S2.** Grid Search Results for the Linear Logistic Regression Classifier | 5 |
| **Table S2.** Results of the hyperparameter tuning for the Random Forest Classifier | 6 |
| **Figure S3.** Grid Search Results for the Random Forest Classifier | 7 |
| **Table S3.** Results of the hyperparameter tuning for the Support Vector Machine Classifier | 8 |
| **Figure S4.** Grid Search Results for the Support Vector Machine Classifier | 10 |
| **Table S4.** Results of the hyperparameter tuning for the Ridge Regression | 11 |
| **Figure S5.** Grid Search Results for the Ridge Regression | 12 |
| **Table S5.** Results of the hyperparameter tuning for the Random Forest Regressor | 13 |
| **Figure S6.** Grid Search Results for the Random Forest Regressor | 14 |
| **Figure S7.** Grid Search Results for the Support Vector Regressor | 15 |
| **Figure S8.** Confusion matrix plots comparing the performance of machine learning classifiers for depression | 16 |
| **Table S6.** Classification performance of machine learning algorithms and dummy baseline models on test set for predicting depression in college students | 17 |
| **Table S7.** Regression performance of machine learning algorithms and dummy baseline models on test set for predicting depression in college students | 18 |
| **Figure S9.** Correlation between actual and predicted values for each machine learning algorithm for the regression task on the test set | 19 |
| **Table S8.** Comparison of the predictive performance of multivariate (all features included) versus univariate (single feature) machine learning models in the classification task | 20 |
| **Table S9.** Comparison of the predictive performance of multivariate (all features included) versus univariate (single feature) machine learning models in the regression task | 21 |

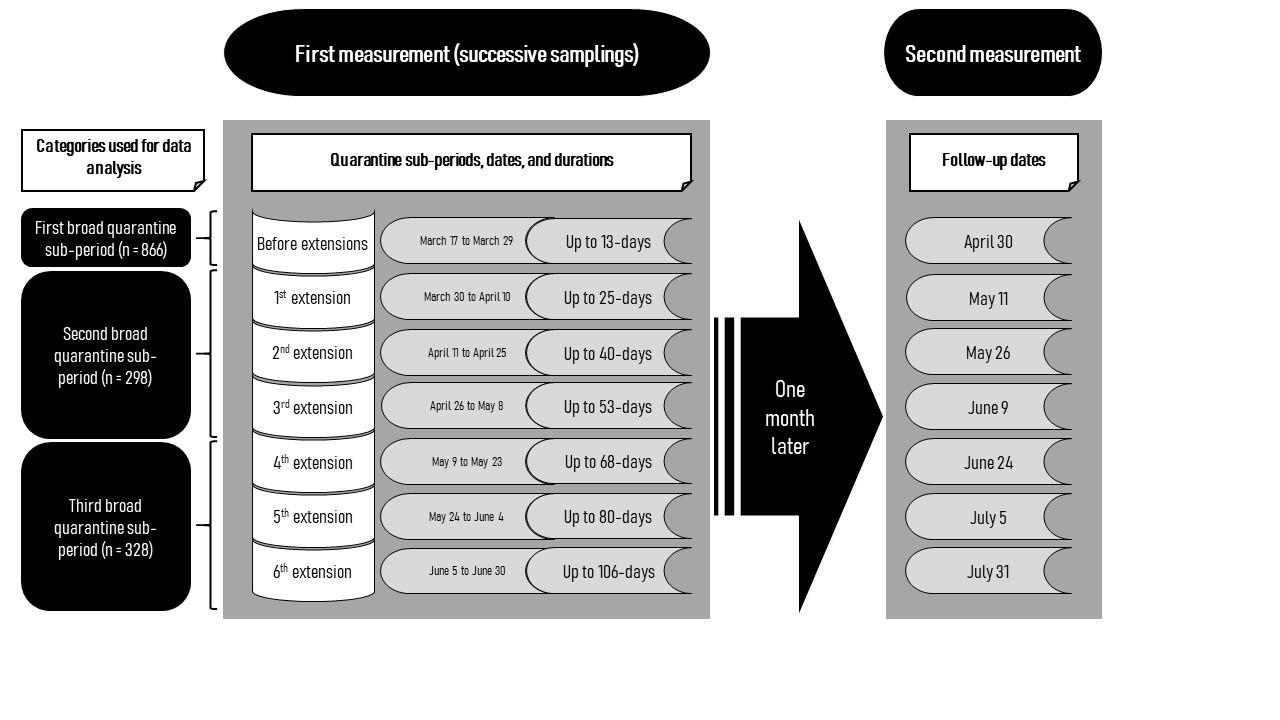

**Figure S1.** *Description of the sampling design*. The first measurement was carried out along successive samplings during the Argentinean quarantine sub-periods. These sub-periods were based on the dates of the Argentinean Governments’ announcements on mandatory quarantine and their extensions. For the first measurement, successive samplings were carried out up to the sixth extension, corresponding to a quarantine of up to 106-days duration. The second measurement was carried out one month later. For data analysis, categories corresponding to three broad quarantine sub-periods were used.

| **Table S1.** Results of the hyperparameter tuning for the Linear Logistic Regression Classifier | | |
| --- | --- | --- |
|  | **Cross-validation mean validation score** | **Hyperparameters** |
| 0 | 0.795241 | C: 0.0001 |
| 1 | 0.797343 | C: 0.001 |
| 2 | 0.797071 | C: 0.01 |
| 3 | **0.798839** | **C: 0.1** |
| 4 | 0.798001 | C: 1 |
| 5 | 0.797562 | C: 10 |
| 6 | 0.797581 | C: 1000 |
| *Note*: The best hyperparameter selected for the linear logistic regression classifier is in bold. C: Regularization parameter. | | |

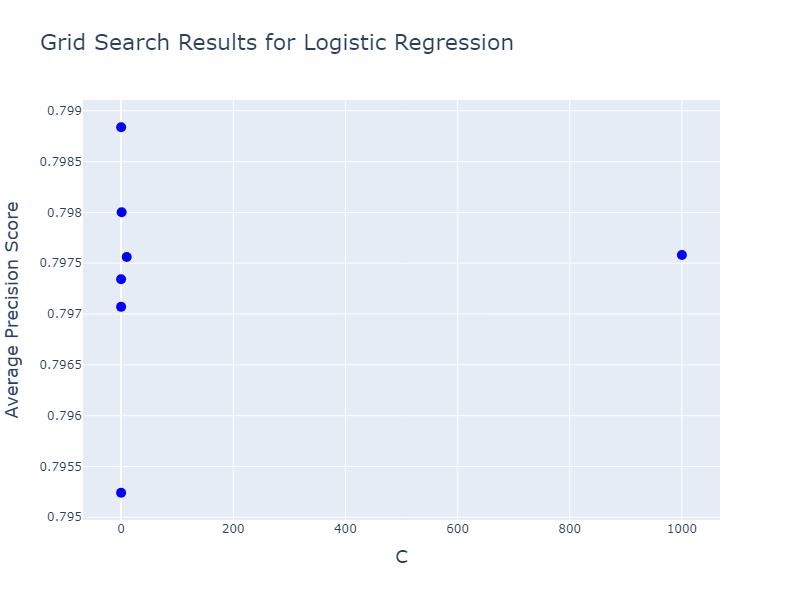

**Figure S2.** *Grid Search Results for the Linear Logistic Regression Classifier*. Scatter plot of the average precision scores for a linear logistic regression model during a grid search. The x-axis represents the values of C used in the hyperparameter search, and the y-axis represents the average precision scores obtained through cross-validation. *Notes*: C: Regularization parameter.

| **Table S2.** Results of the hyperparameter tuning for the Random Forest Classifier | | |
| --- | --- | --- |
|  | **Cross-validation mean validation score** | **Hyperparameters** |
| 0 | 0.748181 | n_estimators: 100 |
| 1 | **0.752826** | **n_estimators: 500** |
| 2 | 0.750381 | n_estimators: 1000 |
| 3 | 0.749035 | n_estimators: 5000 |
| 4 | 0.749684 | n_estimators: 10000 |
| *Note*: The best hyperparameter selected for the random forest classifier is in bold. *Abbreviations*: n_estimators: Number of trees. | | |

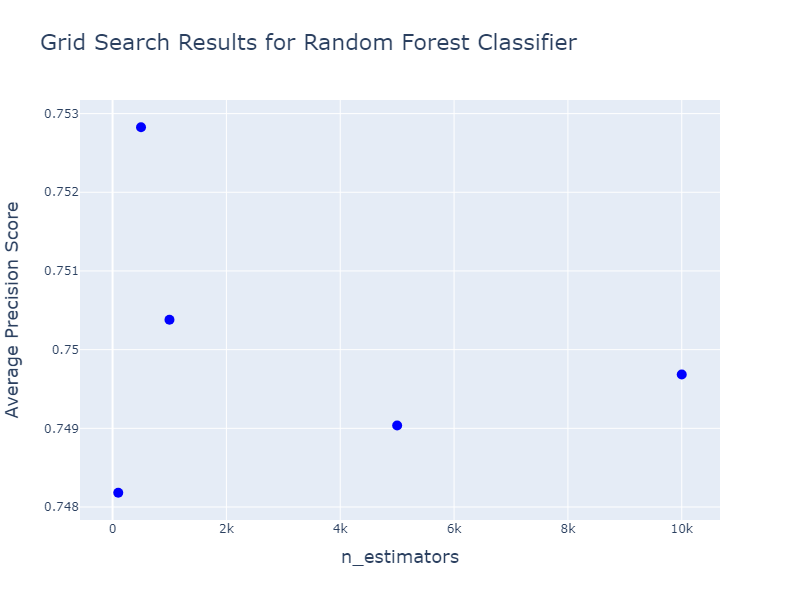

**Figure S3.** *Grid Search Results for the Random Forest Classifier*. Scatter plot of the average precision scores for a random forest classifier model during a grid search. The x-axis represents the number of estimators used in the hyperparameter search, and the y-axis represents the average precision scores obtained through cross-validation. *Notes*: n_estimators: Number of trees.

| **Table S3.** Results of the hyperparameter tuning for the Support Vector Machine Classifier | | |
| --- | --- | --- |
|  | **Cross-validation mean validation score** | **Hyperparameters** |
| 0 | 0.794651 | C: 0.01, gamma: 1e-05, kernel: rbf |
| 1 | 0.794631 | C: 0.01,'gamma: 0.0001, kernel: rbf |
| 2 | 0.794857 | C: 0.01, gamma: 0.001, kernel: rbf |
| 3 | 0.794826 | C: 0.01, gamma: 0.01, kernel: rbf |
| 4 | 0.791338 | C: 0.01, gamma: 0.1, kernel: rbf |
| 5 | 0.741115 | C: 0.01, gamma: 1, kernel: rbf |
| 6 | 0.626249 | C: 0.01, gamma: 10, kernel: rbf |
| 7 | 0.794651 | C: 0.1, gamma: 1e-05, kernel: rbf |
| 8 | 0.794631 | C: 0.1, gamma: 0.0001, kernel: rbf |
| 9 | 0.794725 | C: 0.1, gamma: 0.001, kernel: rbf |
| 10 | 0.796703 | C: 0.1, gamma: 0.01, kernel: rbf |
| 11 | 0.790996 | C: 0.1, gamma: 0.1, kernel: rbf |
| 12 | 0.744310 | C: 0.1, gamma: 1, kernel: rbf |
| 13 | 0.626249 | C: 0.1, gamma: 10, kernel: rbf |
| 14 | 0.794651 | C: 1, gamma: 1e-05, kernel: rbf |
| 15 | 0.794469 | C: 1, gamma: 0.0001, kernel: rbf |
| 16 | 0.796638 | C: 1, gamma: 0.001, kernel: rbf |
| 17 | 0.798668 | C: 1, gamma: 0.01, kernel: rbf |
| 18 | 0.783474 | C: 1, gamma: 0.1, kernel: rbf |
| 19 | 0.697051 | C: 1, gamma: 1, kernel: rbf |
| 20 | 0.605214 | C: 1, gamma: 10, kernel: rbf |
| 21 | 0.794469 | C: 10, gamma: 1e-05, kernel: rbf |
| 22 | 0.795930 | C: 10, gamma: 0.0001, kernel: rbf |
| 23 | 0.800951 | C: 10, gamma: 0.001, kernel: rbf |
| 24 | 0.792662 | C: 10, gamma: 0.01, kernel: rbf |
| 25 | 0.772164 | C: 10, gamma: 0.1, kernel: rbf |
| 26 | 0.624238 | C: 10, gamma: 1, kernel: rbf |
| 27 | 0.564755 | C: 10, gamma: 10, kernel: rbf |
| 28 | 0.795795 | C: 100, gamma: 1e-05, kernel: rbf |
| 29 | 0.801177 | C: 100, gamma: 0.0001, kernel: rbf |
| 30 | 0.795544 | C: 100, gamma: 0.001, kernel: rbf |
| 31 | 0.783134 | C: 100, gamma: 0.01, kernel: rbf |
| 32 | 0.726780 | C: 100, gamma: 0.1, kernel: rbf |
| 33 | 0.579707 | C: 100, gamma: 1, kernel: rbf |
| 34 | 0.563953 | C: 100, gamma: 10, kernel: rbf |
| 35 | 0.800709 | C: 500, gamma: 1e-05, kernel: rbf |
| 36 | 0.797627 | C: 500, gamma: 0.0001, kernel: rbf |
| 37 | 0.791916 | C: 500, gamma: 0.001, kernel: rbf |
| 38 | 0.778532 | C: 500, gamma: 0.01, kernel: rbf |
| 39 | 0.683220 | C: 500, gamma: 0.1, kernel: rbf |
| 40 | 0.572323 | C: 500, gamma: 1, kernel: rbf |
| 41 | 0.563153 | C: 500, gamma: 10, kernel: rbf |
| 42 | **0.801209** | **C: 1000, gamma: 1e-05, kernel: rbf** |
| 43 | 0.797197 | C: 1000, gamma: 0.0001, kernel: rbf |
| 44 | 0.790851 | C: 1000, gamma: 0.001, kernel: rbf |
| 45 | 0.777073 | C: 1000, gamma: 0.01, kernel: rbf |
| 46 | 0.675762 | C: 1000, gamma: 0.1, kernel: rbf |
| 47 | 0.574759 | C: 1000, gamma: 1, kernel: rbf |
| 48 | 0.562591 | C: 1000, gamma: 10, kernel: rbf |
| 49 | 0.800731 | C: 0.01, kernel: linear |
| 50 | 0.797639 | C: 0.1, kernel: linear |
| 51 | 0.796875 | C: 1, kernel: linear |
| 52 | 0.796778 | C: 10, kernel: linear |
| 53 | 0.796884 | C: 100, kernel: linear |
| 54 | 0.797044 | C: 500, kernel: linear |
| 55 | 0.797214 | C: 1000, kernel: linear |
| *Note*: The best hyperparameter combination selected for the support vector machine classifier is in bold. C: Regularization parameter. rbf: Radial basis function. | | |

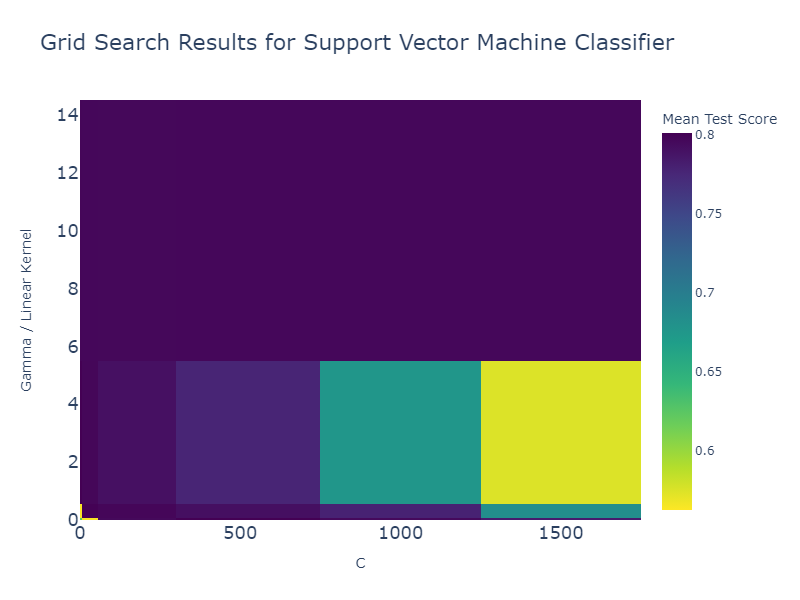

**Figure S4.** *Grid Search Results for the Support Vector Machine Classifier*. Heatmap of the performance of the support vector machine classifier across different hyperparameter values during a grid search. The x-axis represents the regularization parameter, C, while the y-axis corresponds to the kernel parameter, with values specified for the radial basis function (rbf) and linear kernels. The color intensity reflects the mean test score (average precision score), indicating the classifier’s average performance under different configurations. Warmer colors signify higher scores. *Notes*: C: Regularization parameter.

| **Table S4.** Results of the hyperparameter tuning for the Ridge Regression | | |
| --- | --- | --- |
|  | **Cross-validation mean validation score** | **Hyperparameters** |
| 0 | 0.513599 | alpha: 0.0001 |
| 1 | 0.513599 | alpha: 0.001 |
| 2 | 0.513600 | alpha: 0.01 |
| 3 | 0.513605 | alpha: 0.1 |
| 4 | 0.513658 | alpha: 1 |
| 5 | 0.514139 | alpha: 10 |
| 6 | **0.515707** | **alpha: 100** |
| 7 | 0.463237 | alpha: 1000 |
| *Note*: The best hyperparameter selected for the ridge regression is in bold. Alpha: Regularization parameter. | | |

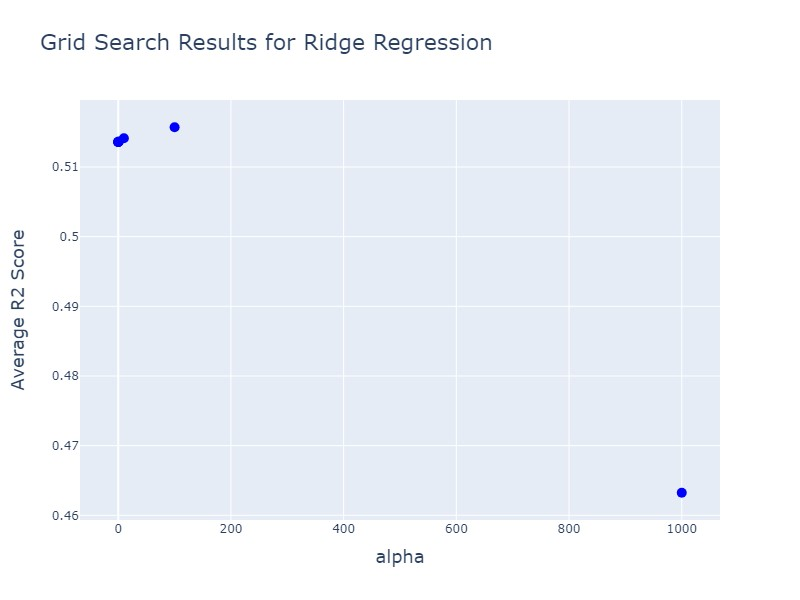

**Figure S5.** *Grid Search Results for the Ridge Regression*. Scatter plot of the average R2 scores for a ridge regression model during a grid search. The x-axis represents the values of the regularization parameter (alpha) used in the hyperparameter search, and the y-axis represents the average R2 scores obtained through cross-validation. *Notes*: Alpha: regularization parameter. R2: R-squared.

| **Table S5.** Results of the hyperparameter tuning for the Random Forest Regressor | | |
| --- | --- | --- |
|  | **Cross-validation mean validation score** | **Hyperparameters** |
| 0 | 0.443716 | n_estimators: 100 |
| 1 | 0.443881 | n_estimators: 500 |
| 2 | 0.444682 | n_estimators: 1000 |
| 3 | **0.445658** | **n_estimators: 5000** |
| 4 | 0.445238 | n_estimators: 10000 |
| *Note*: The best hyperparameter selected for the random forest regressor is in bold. *Notes*: n_estimators: Number of trees. | | |

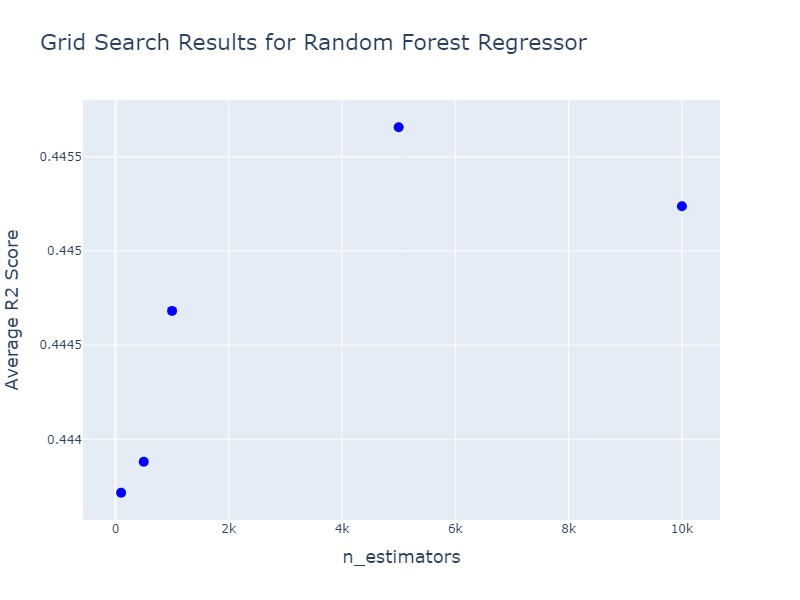

**Figure S6.** *Grid Search Results for the Random Forest Regressor*. Scatter plot of the results of the average R2 scores for a random forest regressor model during a grid search. The x-axis represents the number of estimators used in the hyperparameter search, and the y-axis represents the average R2 scores obtained through cross-validation. *Notes*: n_estimators: Number of trees. R2: R-squared.

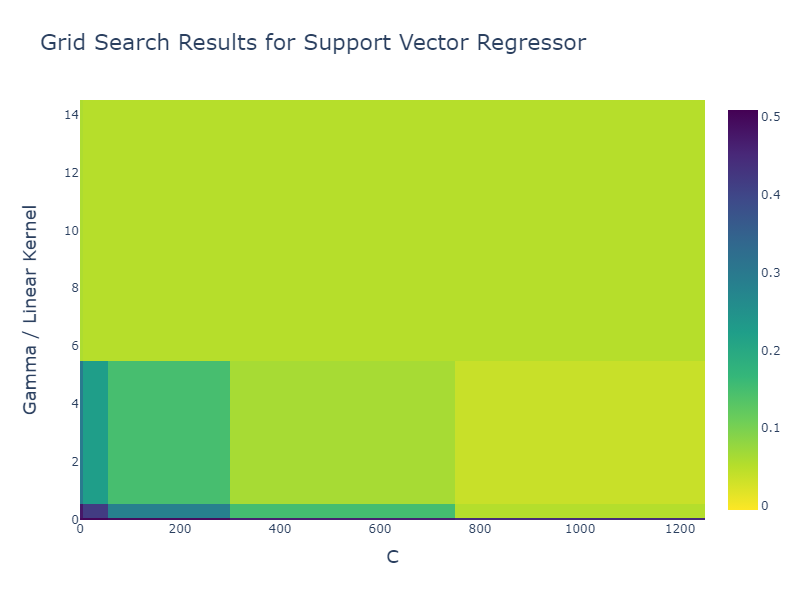

**Figure S7.** *Grid Search Results for the Support Vector Regressor (SVR)*. Heatmap of the performance of the support vector machine regressor across different hyperparameter values during a grid search. The x-axis represents the values of the regularization parameter, C. The y-axis displays the values of the hyperparameter Gamma for the radial basis function (rbf) kernel, or indicates linear for the linear kernel. The color of each cell corresponds to the mean R2 score achieved during cross-validation for the respective combination of C, Gamma, and kernel type. Darker colors represent higher mean test scores. *Notes*: C: Regularization parameter. R2: R-squared.

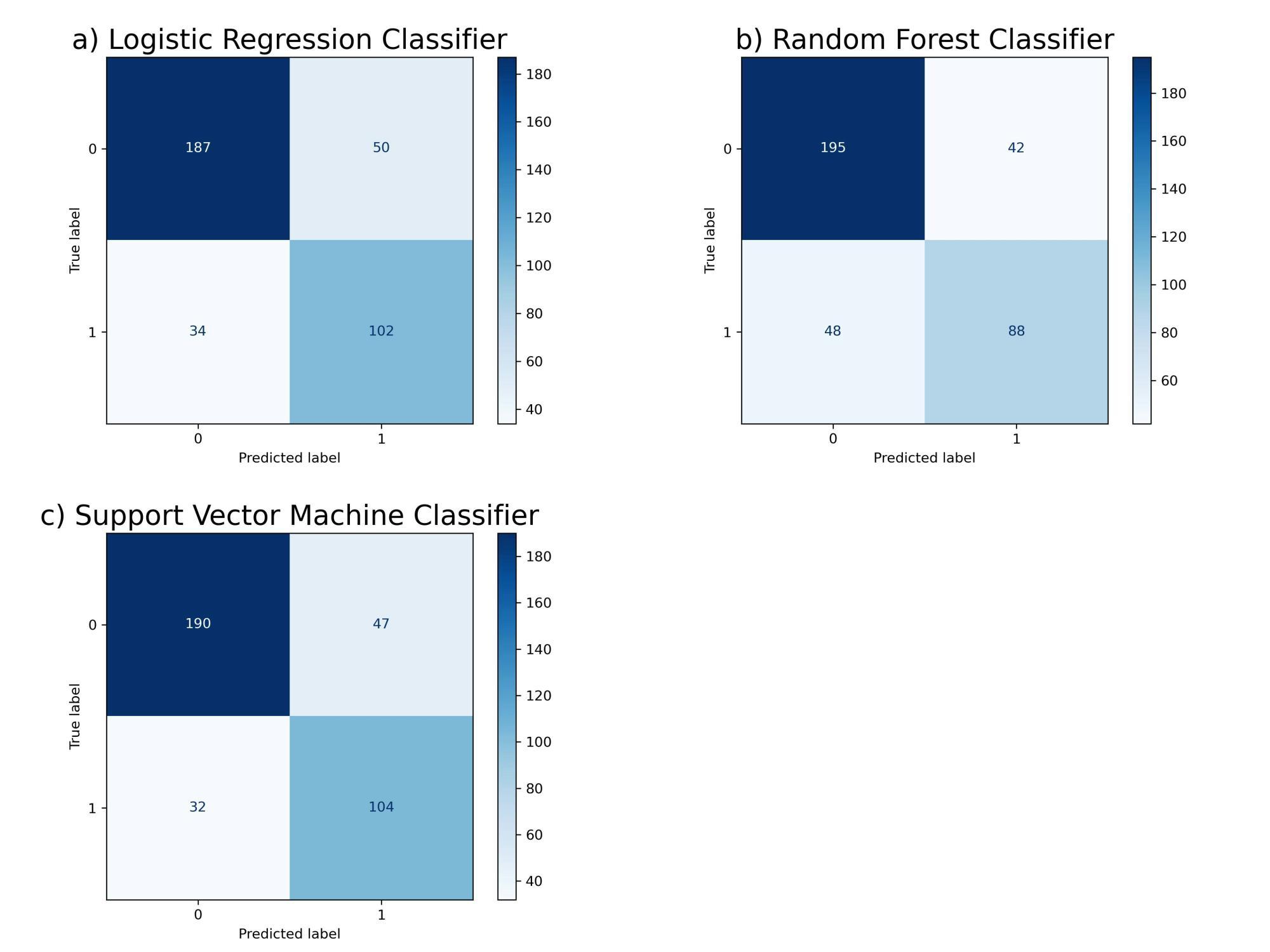

**Figure S8.** *Confusion matrix plots comparing the performance of machine learning classifiers for depression*. Each plot shows the number of correct and incorrect predictions of depression labels (1 = depressed, 0 = non-depressed). Darker color indicates a higher number of correct predictions. **a**) Linear Logistic Regression Classifier. **b**) Random Forest Classifier. **c**) Support Vector Machine Classifier.

| **Table S6.** Classification performance of machine learning algorithms and dummy baseline models on the test set for predicting depression in college students | | | | | | | |
| --- | --- | --- | --- | --- | --- | --- | --- |
| **Dummy (baselines) and classification models** | **Performance metrics for baseline and machine learning classification models on the test set** | | | | | | |
|  | **AUPRC** | **AUROC** | **Balanced accuracy** | **1 - Brier loss** | **F1 score** | **Precision** | **Recall** |
| Uniform Random Baseline | **0.36**  (0.33, 0.40) | **0.50**  (0.50, 0.50) | **0.50**  (0.45, 0.55) | **0.75**  (0.75, 0.75) | **0.43**  (0.37, 0.49) | **0.36**  (0.30, 0.43) | **0.52**  (0.46, 0.58) |
| Most Frequent Baseline | **0.36**  (0.33, 0.40) | **0.50**  (0.50, 0.50) | **0.50**  (0.50, 0.50) | **0.64**  (0.60, 0.67) | **0.00**  (0.00, 0.00) | **1.00**  (1.00, 1.00) | **0.00**  (0.00, 0.00) |
| Stratified Random Baseline | **0.36**  (0.32, 0.42) | **0.50**  (0.44, 0.55) | **0.50**  (0.44, 0.55) | **0.54**  (0.49, 0.58) | **0.36**  (0.30, 0.43) | **0.36**  (0.29, 0.45) | **0.36**  (0.29, 0.42) |
| Linear Logistic Regression | **0.76**  (0.69, 0.81) | **0.85**  (0.80, 0.88) | **0.77**  (0.72, 0.80) | **0.84**  (0.82, 0.86) | **0.71**  (0.65, 0.75) | **0.67**  (0.60, 0.73) | **0.75**  (0.67, 0.82) |
| Random Forest Classifier | **0.72**  (0.65, 0.80) | **0.82**  (0.78, 0.86) | **0.74**  (0.70, 0.77) | **0.84**  (0.81, 0.86) | **0.66**  (0.59, 0.72) | **0.68**  (0.62, 0.76) | **0.65**  (0.58, 0.73) |
| Support Vector Machine Classifier | **0.76**  (0.69, 0.81) | **0.85**  (0.80, 0.88) | **0.78**  (0.74, 0.82) | **0.85**  (0.83, 0.86) | **0.72**  (0.67, 0.76) | **0.69**  (0.63, 0.76) | **0.76**  (0.70, 0.83) |
| *Notes*: For the baselines and machine learning classification models in the test set, the mean scores obtained from model predictions across 100 simulated samplings using the bootstrap method are indicated in bold, with the lower and upper limits of 95% confidence interval in parentheses. The *1 - Brier loss* is presented as the inverse of the Brier score to ensure higher values consistently denote better performance across all methods. Original values of the Brier loss score are as follows: Uniform random baseline 0.25 (0.25, 0.25), Most frequent baseline 0.36 (0.33, 0.40), Stratified random baseline 0.46 (0.42, 0.51), Linear logistic regression 0.16 (0.14, 0.18), Random forest classifier 0.16 (0.14, 0.19), and SVM classifier 0.15 (0.14, 0.17). *Abbreviations*: AUPRC: Area Under the Precision-Recall Curve. AUROC: Area Under the Receiver Operating Characteristic Curve. | | | | | | | |

| **Table S7.** Regression performance of machine learning algorithms and dummy baseline models on the test set for predicting depression in college students | | | |
| --- | --- | --- | --- |
| **Dummy (baselines) and regression models** | **Performance metrics for baseline and machine learning regression models on the test set** | | |
|  | **R2** | **Mean Absolute Error** | **Mean Squared Error** |
| Randomly Shuffled Baseline | **-1.02**  (-1.33, -0.78) | **1.08**  (1.01, 1.17) | **1.85**  (1.65, 2.13) |
| Mean Baseline | **-0.003**  (-0.01, -0.00001) | **0.78**  (0.72, 0.85) | **0.92**  (0.77, 1.10) |
| Median Baseline | **-0.003**  (-0.01, -0.00002) | **0.78**  (0.72, 0.85) | **0.92**  (0.77, 1.10) |
| Ridge Regression | **0.56**  (0.45, 0.63) | **0.50**  (0.46, 0.54) | **0.40**  (0.34, 0.47) |
| Random Forest Regressor | **0.49**  (0.38, 0.57) | **0.53**  (0.48, 0.57) | **0.46**  (0.40, 0.55) |
| Support Vector Regressor | **0.56**  (0.45, 0.64) | **0.50**  (0.46, 0.53) | **0.40**  (0.33, 0.47) |
| *Notes*: For the baselines and machine learning regression models in the test set, the mean scores obtained from model predictions across 100 simulated samplings using the bootstrap method are indicated in bold, with the lower and upper limits of 95% confidence interval in parentheses. *Abbreviations*: R2: R-squared. | | | |

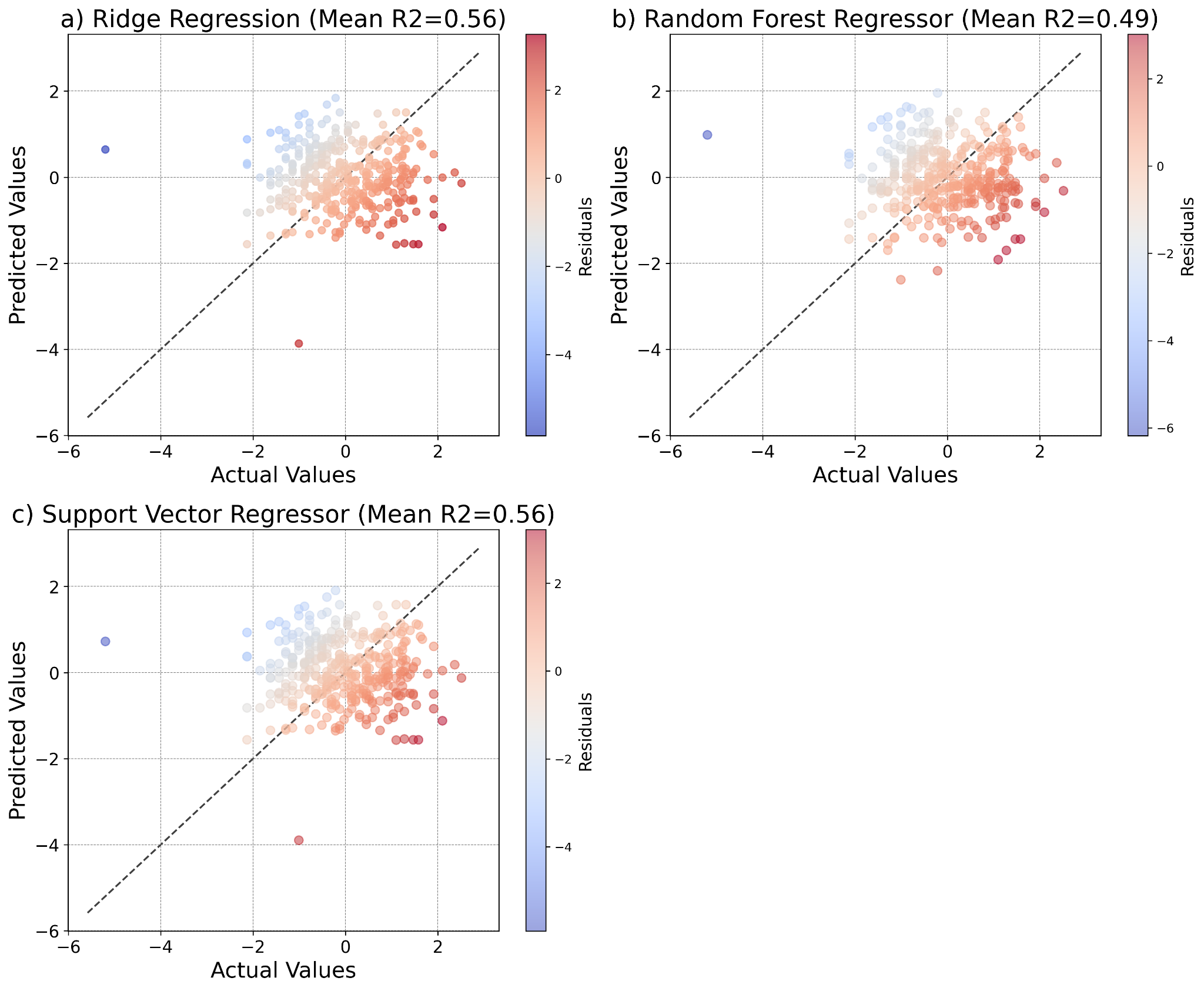

**Figure S9.** *Correlation between actual and predicted values for each machine learning algorithm for the regression task on the test set*. Plots show actual and predicted depression scores of three regression models for individual study participants alongside color-coded residual values. **a**) Ridge Regression. **b**) Random Forest Regressor. **c**) Support Vector Regressor.

| **Table S8.** Comparison of the predictive performance of multivariate (all features included) versus univariate (single feature) machine learning models in the classification task | | | | |
| --- | --- | --- | --- | --- |
|  |  | **Mean AUPRC scores (95% CI lower, upper)** | | |
|  | **Features** | **Linear logistic regression** | **Random forest classifier** | **Support vector machine classifier** |
| Multivariate model (includes all features) | | 0.76 (0.69, 0.81) | 0.72 (0.65, 0.80) | 0.76 (0.69, 0.81) |
| Univariate models | Depression T1 | 0.72 (0.64, 0.79) | 0.73 (0.67, 0.78) | 0.72 (0.64, 0.79) |
|  | Anxiety | 0.71 (0.64, 0.78) | 0.68 (0.59, 0.75) | 0.71 (0.64, 0.78) |
|  | Quarantine sub-period | 0.37 (0.33, 0.42) | 0.37 (0.33, 0.42) | 0.41 (0.35, 0.47) |
|  | Sex | 0.38 (0.34, 0.42) | 0.38 (0.34, 0.42) | 0.38 (0.34, 0.42) |
|  | Age | 0.43 (0.37, 0.50) | 0.40 (0.35, 0.49) | 0.43 (0.37, 0.50) |
|  | Mental disorder history | 0.46 (0.40, 0.52) | 0.46 (0.40, 0.52) | 0.46 (0.40, 0.52) |
|  | Suicidal behavior history | 0.46 (0.39, 0.52) | 0.46 (0.39, 0.52) | 0.46 (0.39, 0.52) |
| *Abbreviations*: AUPRC: Area Under the Precision-Recall Curve. CI: Confidence intervals. T1: First measurement or measurement at time one. | | | | |

| **Table S9.** Comparison of the predictive performance of multivariate (all features included) versus univariate (single feature) machine learning models in the regression task | | | | | |
| --- | --- | --- | --- | --- | --- |
|  | |  | **Mean R2 scores (95% CI lower, upper)** | | |
|  |  | **Features** | **Ridge regression** | **Random forest regressor** | **Support vector regressor** |
| Multivariate model (includes all features) | | | 0.56 (0.44, 0.64) | 0.49 (0.38, 0.57) | 0.56 (0.45, 0.64) |
| Univariate models | Depression T1 | | 0.48 (0.39, 0.57) | 0.43 (0.26, 0.56) | 0.33 (0.29, 0.38) |
|  | Anxiety | | 0.39 (0.30, 0.47) | 0.37 (0.24, 0.45) | 0.27 (0.22, 0.32) |
|  | Quarantine sub-period | | -0.00 (-0.02, 0.00) | -0.00 (-0.02, 0.00) | -0.04 (-0.08, -0.01) |
|  | Sex | | 0.01 (-0.02, 0.03) | 0.01 (-0.02, 0.03) | -0.03 (-0.08, -0.01) |
|  | Age | | 0.03 (0.00, 0.05) | 0.00 (-0.05, 0.04) | -0.01 (-0.05, 0.01) |
|  | Mental disorder history | | 0.03 (-0.02, 0.07) | 0.03 (-0.02, 0.07) | -0.01 (-0.06, 0.01) |
|  | Suicidal behavior history | | 0.13 (0.07, 0.20) | 0.13 (0.07, 0.20) | 0.02 (-0.02, 0.05) |
| *Abbreviations*: R2: R-squared. CI: Confidence intervals. T1: First measurement or measurement at time one. | | | | | |
